# COVID-19 and Vitamin D (Co-VIVID Study): a systematic review and meta-analysis of randomized controlled trials

**DOI:** 10.1101/2021.08.22.21262216

**Authors:** Seshadri Reddy Varikasuvu, Balachandar Thangappazham, Hemanth Raj

## Abstract

**Background:** Vitamin D levels have been reported to be associated with COVID-19 susceptibility, severity and mortality events.. We performed a meta-analysis of randomized controlled trials (RCTs) to evaluate the use of vitamin D intervention on COVID-19 outcomes.

**Methods:** Literature search was conducted using PubMed, Cochrane library, and ClinicalTrials.gov databases (latest search on August 5, 2021). We included RCTs reporting the use of vitamin D intervention to control/placebo group in COVID-19. Two independent researchers did literature search, abstracted data, and the risk of bias assessment.

**Results:** A total of 6 RCTs with 551 COVID-19 patients were included. The overall collective evidence pooling all the outcomes across all RCTs indicated the beneficial use of vitamin D intervention in COVID-19 (relative risk, RR = 0.60, 95% CI 0.40 to 0.92, Z=2.33, p=0.02, I^2^ = 48%). However, no statistical significance was observed for individual outcomes of ICU care (RR = 0.11, 95% CI 0.15 to 1.30, Z=1.48, p=0.14, I^2^ = 66%) and mortality (RR = 0.78, 95% CI 0.25 to 2.40, Z=0.66, p=0.02, I^2^ = 33%), though decreased rates were noted. The rates of RT-CR positivity was significantly decreased in the intervention group as compared to the non-vitamin D groups (RR = 0.46, 95% CI 0.24 to 0.89, Z=2.31, p=0.02, I^2^ = 0%).

**Conclusion:** COVID-19 patients supplemented with vitamin D are more likely to demonstrate fewer rates of ICU admission, mortality events and RT-PCR positivity. However, no statistical significance has been achieved for individual outcomes of ICU and deaths. More RCTs and completion of ongoing trials largely needed to precisely establish the association between vitamin D use and COVID-19.

## Introduction

Since December 2019, millions have infected with severe acute respiratory syndrome associated with coronavirus-2 (SARS-CoV-2) causing Coronavirus disease 2019 (COVID-19), a global pandemic the World Health Organisation (WHO). The COVID-19 symptoms range from mildly symptomatic to moderate, severe to critical with patients needing hospitalization and intensive care unit (ICU) admissions. As of 5^th^ August 2021 and WHO, there have been 200,174,883 confirmed COVID-19 cases, including 4,255,892 deaths [1]. Multiple risk factors in the form of age, comorbidities, exaggerated immune response in the form of cytokine storm, oxidative stress, activation of pro-coagulation factors and severe inflammation contribute to the disease progression [2].

It has been documented that vitamin D deficiency is associated with severity of viral infections such as influenza [3]. Recent evidence shows the potential of vitamin D to affect SARS-CoV-2 gene expression and alleviate infection upon binding to the vitamin D response element [4,5]. Vitamin D regulates the rennin-angiotensin system and expression of angiotensin converting enzyme 2 (ACE2), and its receptor that mediates SARS-CoV-2 infection. Further, vitamin D is known to exert immuno-modulatory effects in innate and adaptive immune responses, induces the production of antimicrobial proteins and could act as anti-inflammatory agent [4,6,7].

Despite vaccination rollouts, much focus has been documented on additional preventive measures such as using vitamin D supplementation to be promising in COVID-19 [7,8]. While strong observational evidence [9–11] indicate the association of low vitamin D levels to the COVID-19 susceptibility, severity and mortality outcomes, the beneficial use of vitamin D supplements in COVID-19 has been reported in some non-randomized observational cohorts [12,13]. However, there is still a scarcity of information through randomized controlled trials (RCTs) on the use of vitamin D supplementation in COVID-19 patients. With many of the trials in the ongoing stage, there is a greater need for supportive evidence through meta-analysis of available RCTs [14–19]. Therefore, our objective of this study was to evaluate the effect of vitamin D intervention in relationship to several COVID-19 outcomes reported in all available RCTs.

## Material and Methods

This meta-analysis was conducted and reported according to the Preferred Reporting Items for Systematic Reviews and Meta-Analysis (PRISMA) [20]. The protocol was registered at PROSPERO: CRD42021271461.

### Literature search and study selection

The literature search was conducted with no language restrictions using PubMed/MEDLINE, Cochrane library, EMBASE, SCOPUS, Science Direct, and ClinicalTrials.gov from inception to August 5, 2021. The search strategy included both the MeSH and broad text-word search terms: (“vitamin D” (MeSH Terms) OR “vitamin D” (All Fields) OR “ergocalciferols” (MeSH Terms) OR “ergocalciferols” (All Fields)) AND (“SARS-CoV-2 OR COVID-19” (MeSH Terms) OR “SARS-CoV-2 OR COVID-19” (All Fields)). The other terms used for vitamin D were 25-Hydroxyvitamin D, 25-Hydroxycholecalciferol, calcidiol, 1.25-dihydroxyvitamin D3, Calcifediol, and Calcitriol. The other terms used for SARS-CoV-2/COVID-19 are Coronavirus and 2019-nCoV Disease. The bibliographies of published articles were manually hand-searched for additional studies.

The inclusion criteria were: (1) RCTs comparing supplementation of vitamin D to placebo/control; (2) RCTs reporting the use of vitamin D supplementation on one or more of the following; COVID-19 severity, ICU care, mortality events, seropositivity and RT-PCR positivity or any other adverse events. No prespecified limitations applied for dose or type of vitamin D and follow-up durations. The exclusion criteria were: (1) studies with no control/comparator group; (2) study types other than RCTs such as observational studies and trial-protocols. In case of duplicate articles, only a recent report with all relevant information was included. All the relevant RCTs were screened at the title, abstract and full-text levels for their suitability in this systematic review and meta-analysis.

### Data extraction and risk of bias assessment

The information extracted from eligible RCTs include: first author names, study country and setting, sample sizes, randomization, blinding, vitamin D form and dose, follow-up details, number of events for study outcomes (severity, ICU care, mortality, seropositivity and RT-PCR positivity) in treatment and comparator groups, and other study characteristics. Two investigators (S.R.V. and B.T.) independently assessed the potential risks of bias of the RCTs using the Cochrane Risk of Bias Tool [21]. Two authors (S.R.V. and B.T.) have independently performed the literature search, study selection and assessment. Any discrepancies were resolved upon discussion with a third investigator (H.R.). When required, the corresponding authors of respective articles were contacted through e-mail to obtain data/clarification.

### Data analysis

For this meta-analysis of RCTs, we reported the effect sizes as risk ratio (RR) for the number of events on the outcomes such as severity, ICU admissions, mortality, seropositivity and RT-PCR positivity in treated and control groups. We reported RR values with their 95% confidence intervals (CI) using Mantel-Haenszel analysis method and random-effects model. The overall effect size for RR was presented Z-score. A Z-score with a *p* value of <0.05 was considered statistically significant. The between-study heterogeneity was examined by the I^2^ statistics and the values >50% were considered to indicate a high degree of heterogeneity [22]. We examined the funnel plot asymmetry for publication bias followed by Begg and Egger’s tests.

### Sub-group and sensitivity analysis

We conducted sub-group analysis based on vitamin D form, vitamin D deficient studies, single or multi-centric trials, and double-blinded status. We also performed a one-study leave-out sensitivity analysis for individual outcomes by excluding one trial at a time and by repeating the analysis. The meta-regression analysis was not possible due to the small number of available trials.

## Results

We reviewed 755 articles for eligibility, 6 RCTs [14–19] comprising 551 COVID-19 patients were selected for final analysis (Fig. 1).

**Fig.1.**
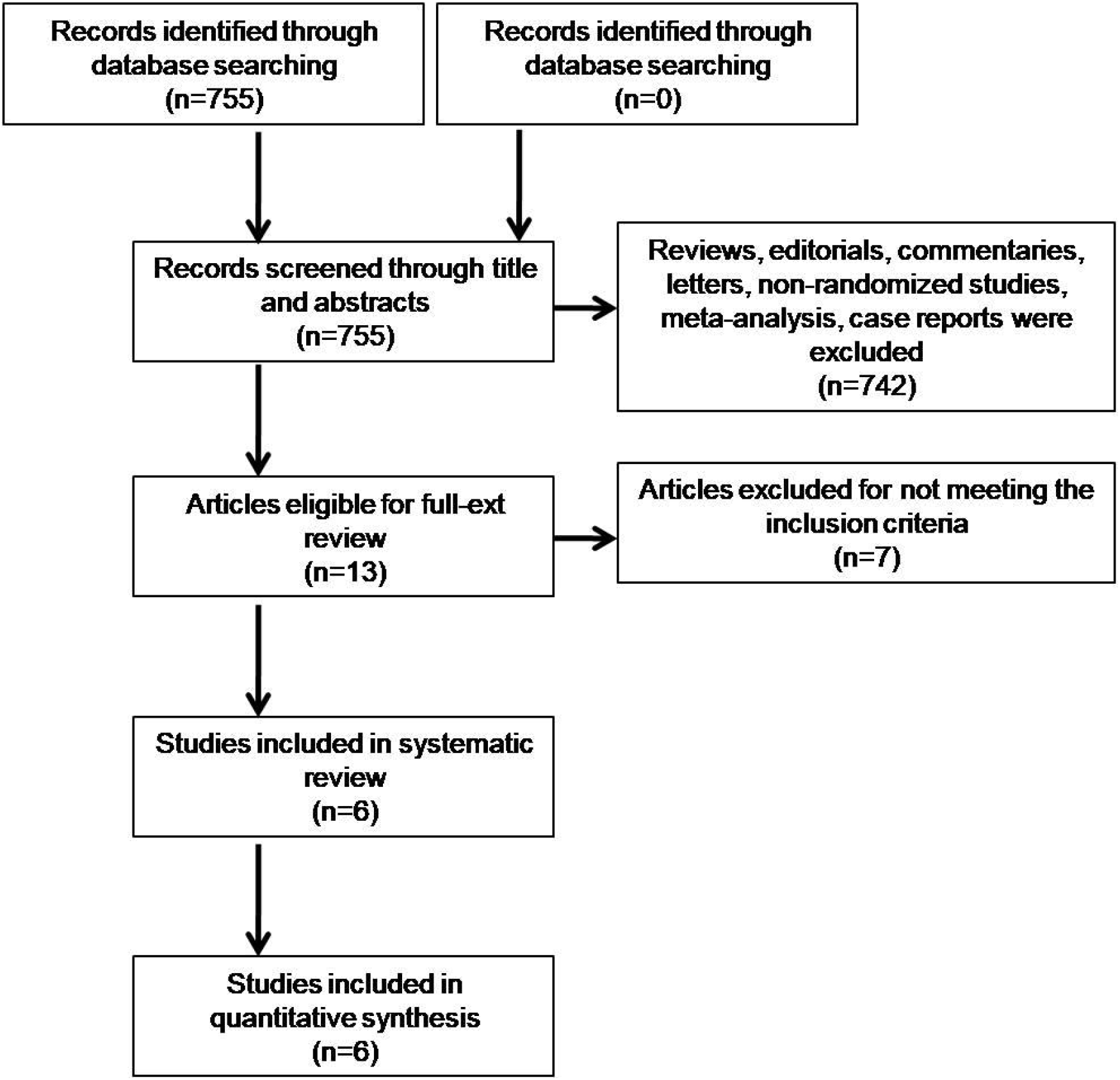
Literature search results

While all the studies enrolled participants aged >18 years with mean age in individual studies range from 36 to 56 years, the proportion of men varied from 44 to 59%. The symptoms of COVID-19 patients diagnosed by RT-PCR (viral RNA) or ELISA and/or radiographic testing varied across the individual studies (mild-moderate-severe). The criteria for inclusion and exclusion, varied study settings, participant characteristics, number of participants with preexisting comorbidities and treatment strategies, vitamin D form, dosage, reported outcomes and other study characteristics are presented in Table 1.

**Table 1.**
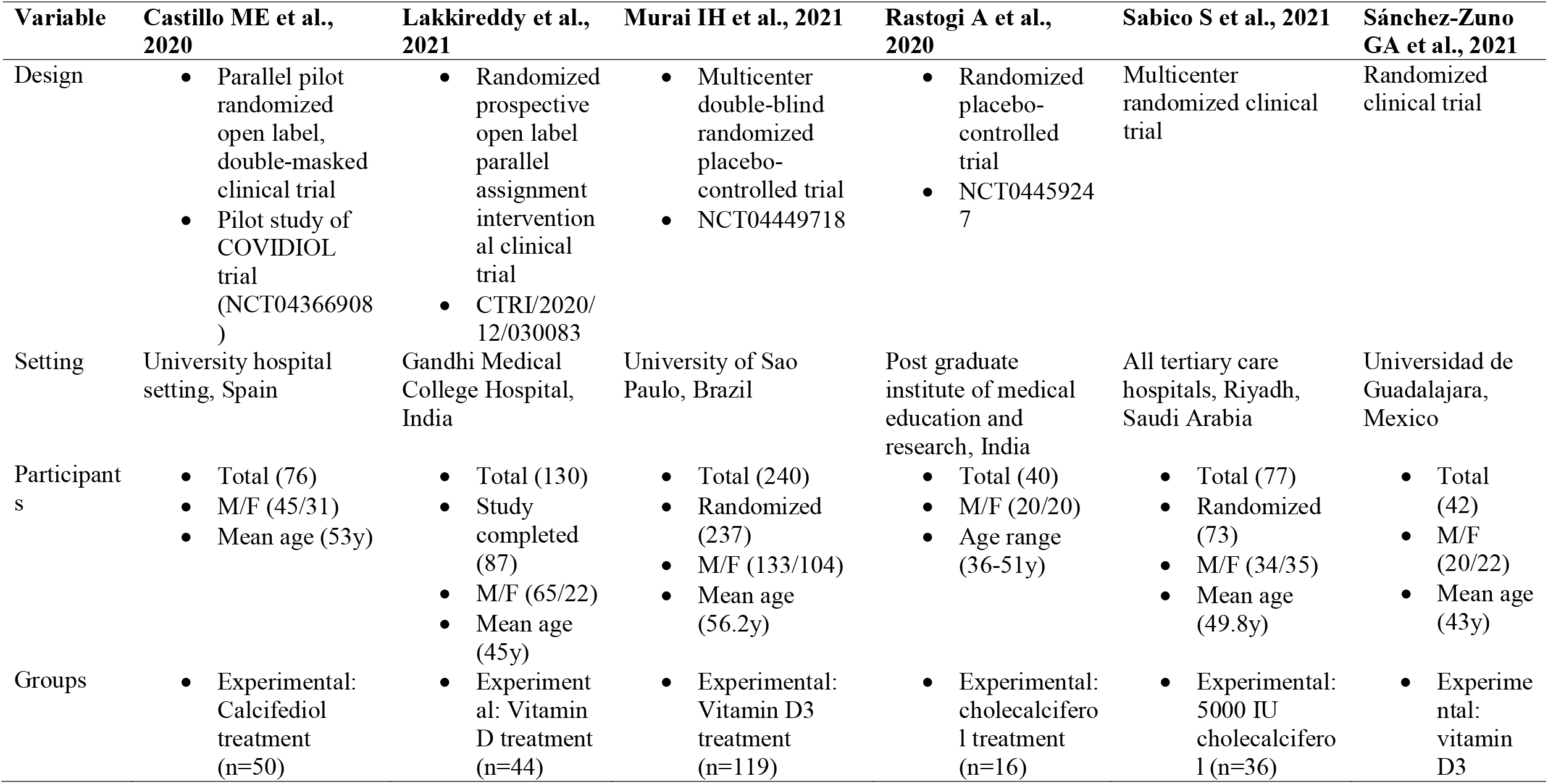
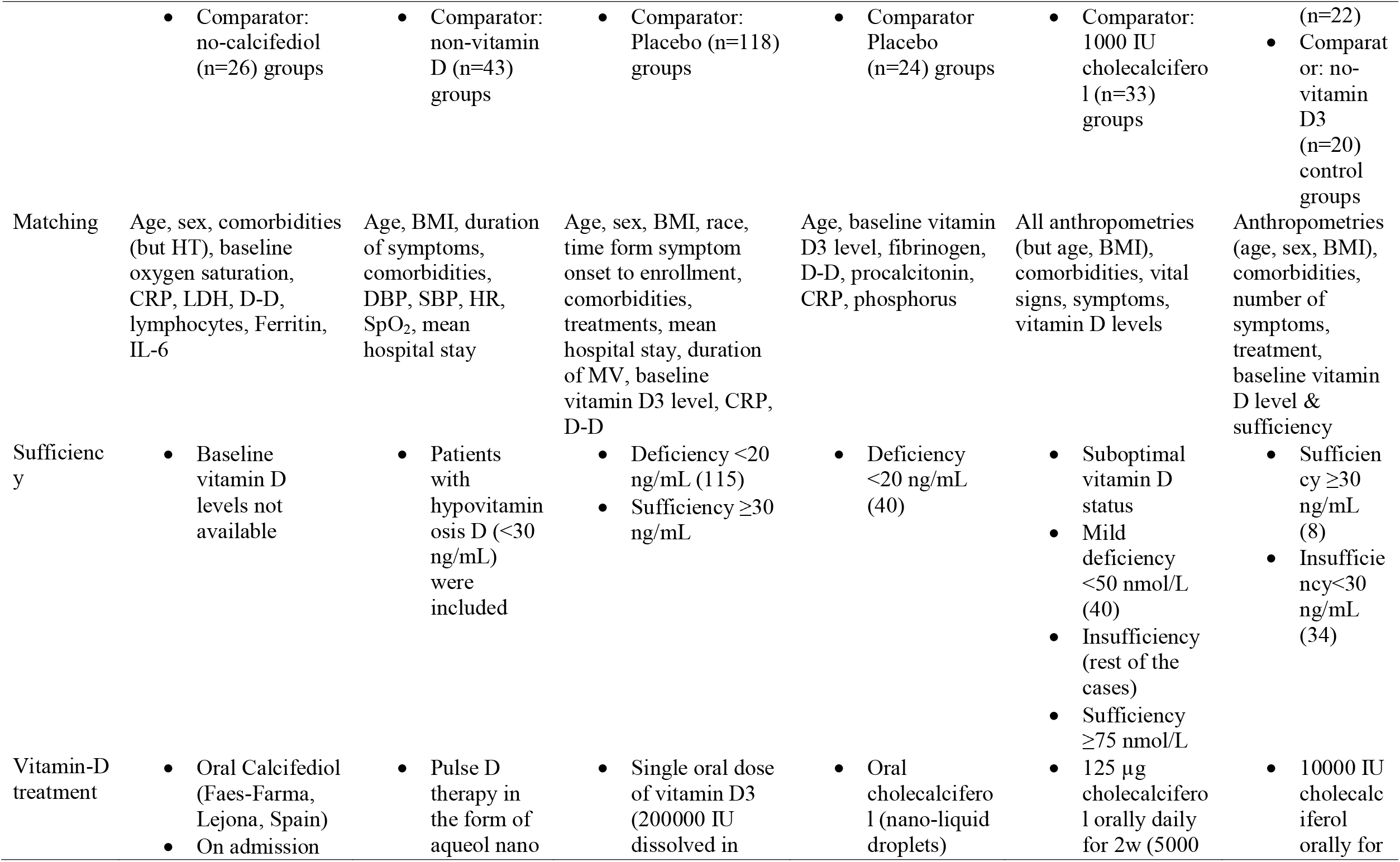
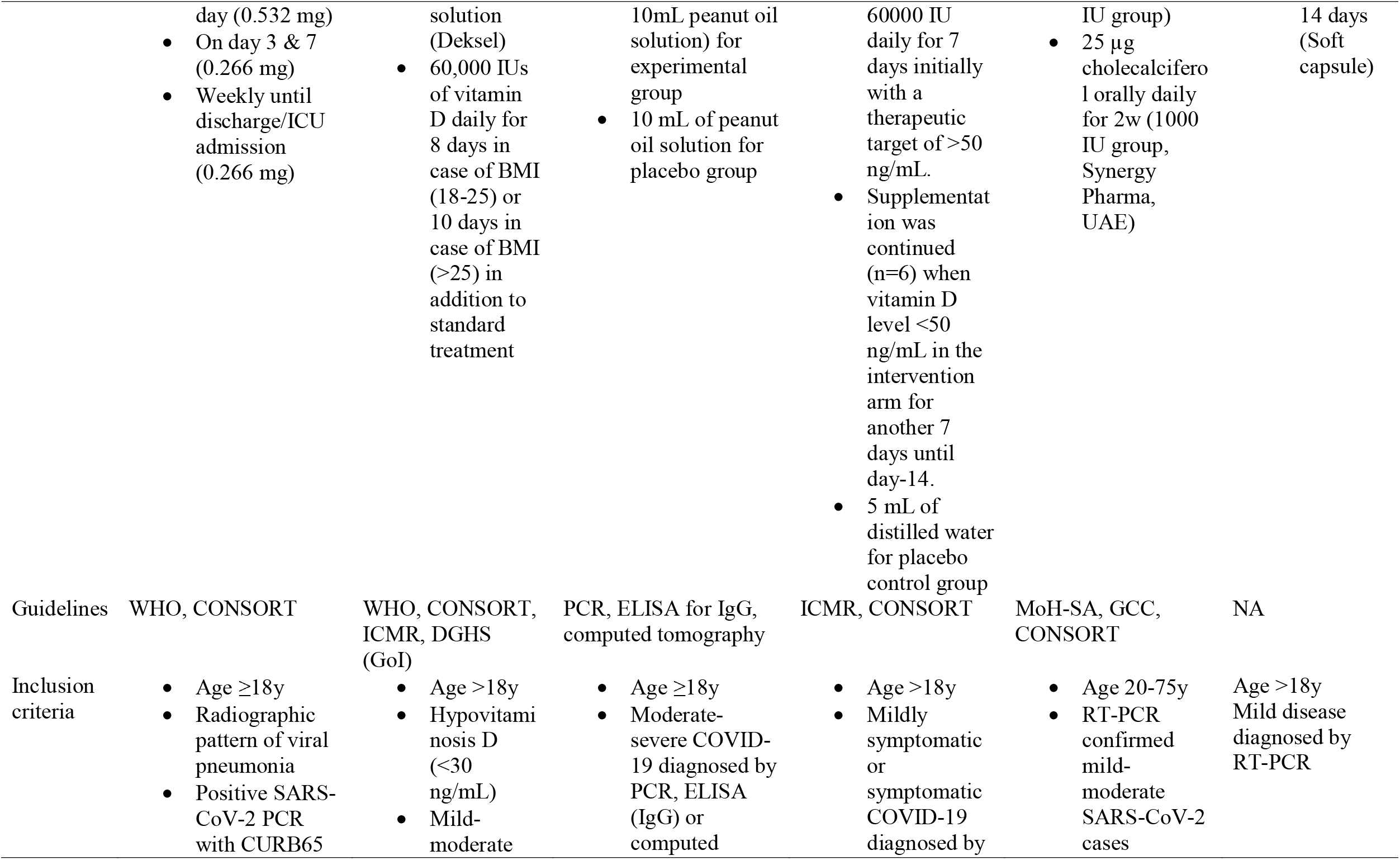
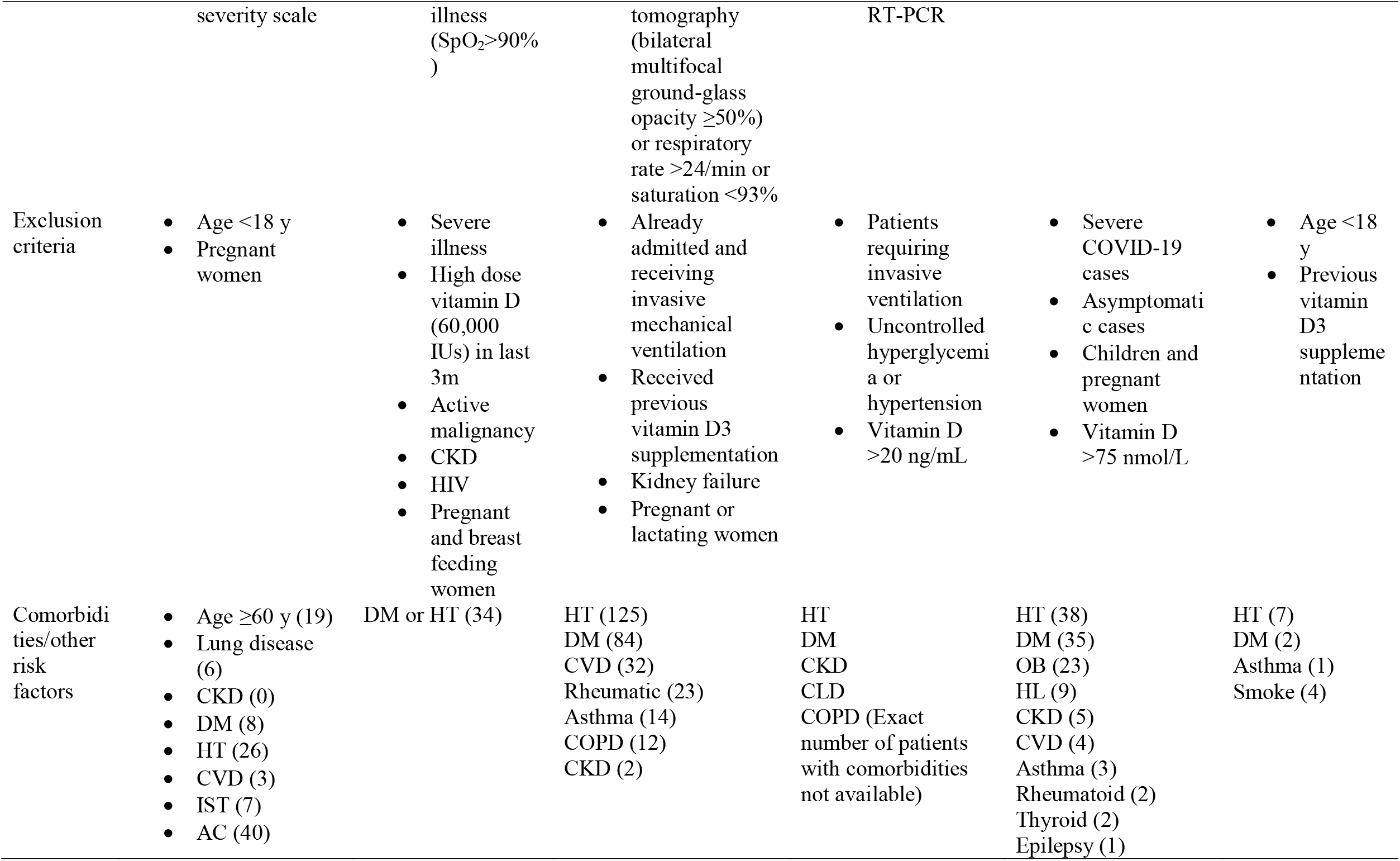
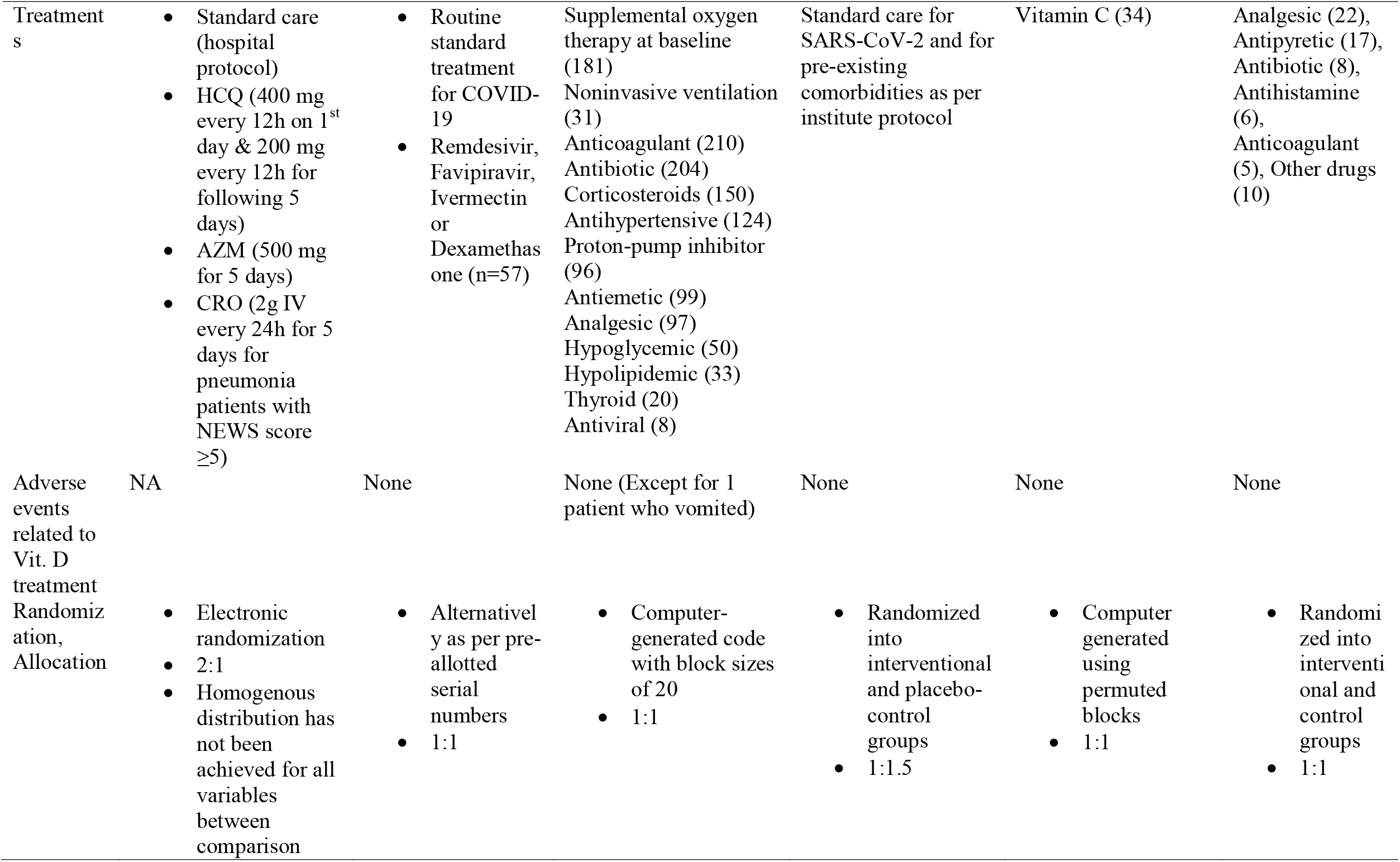
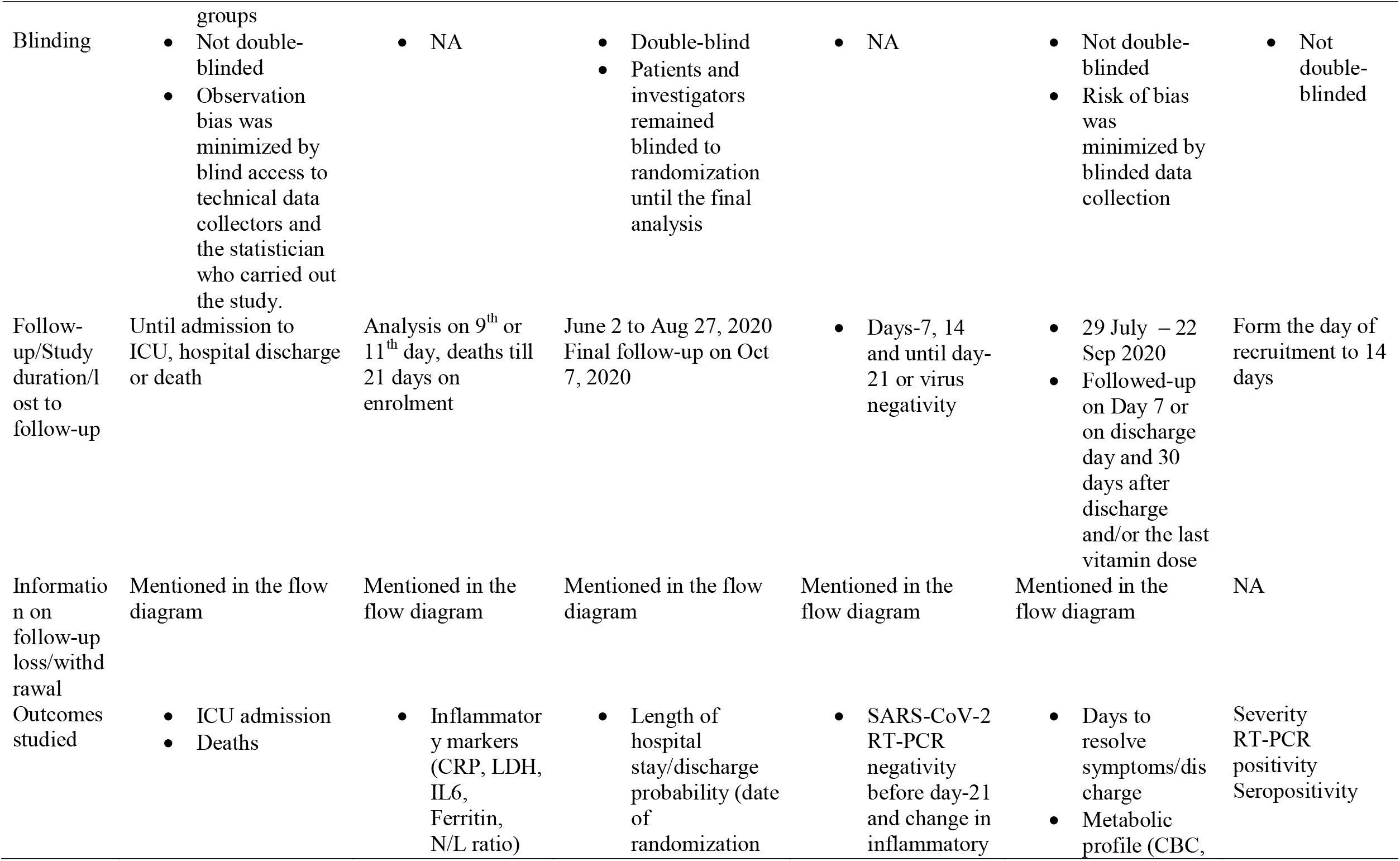
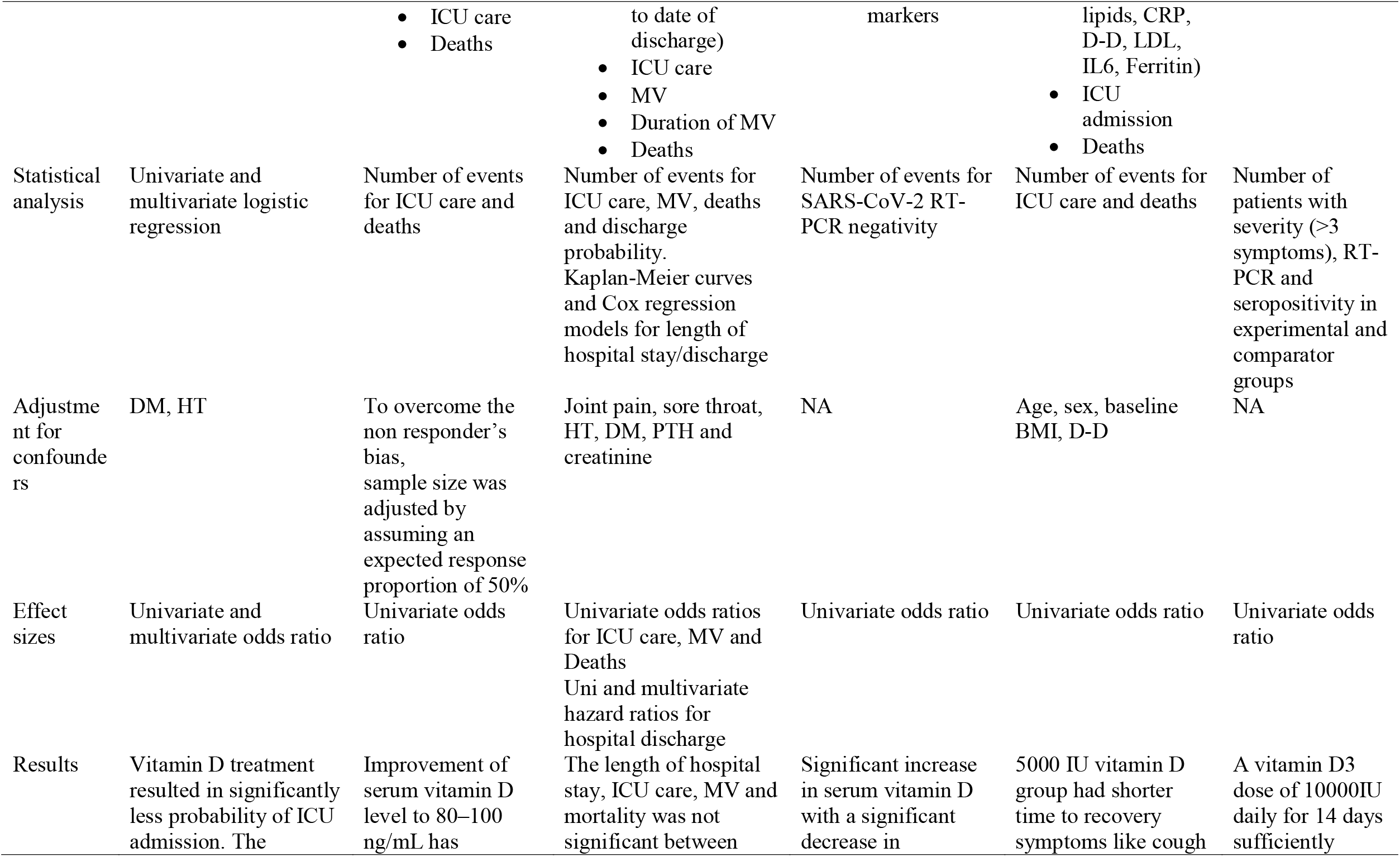
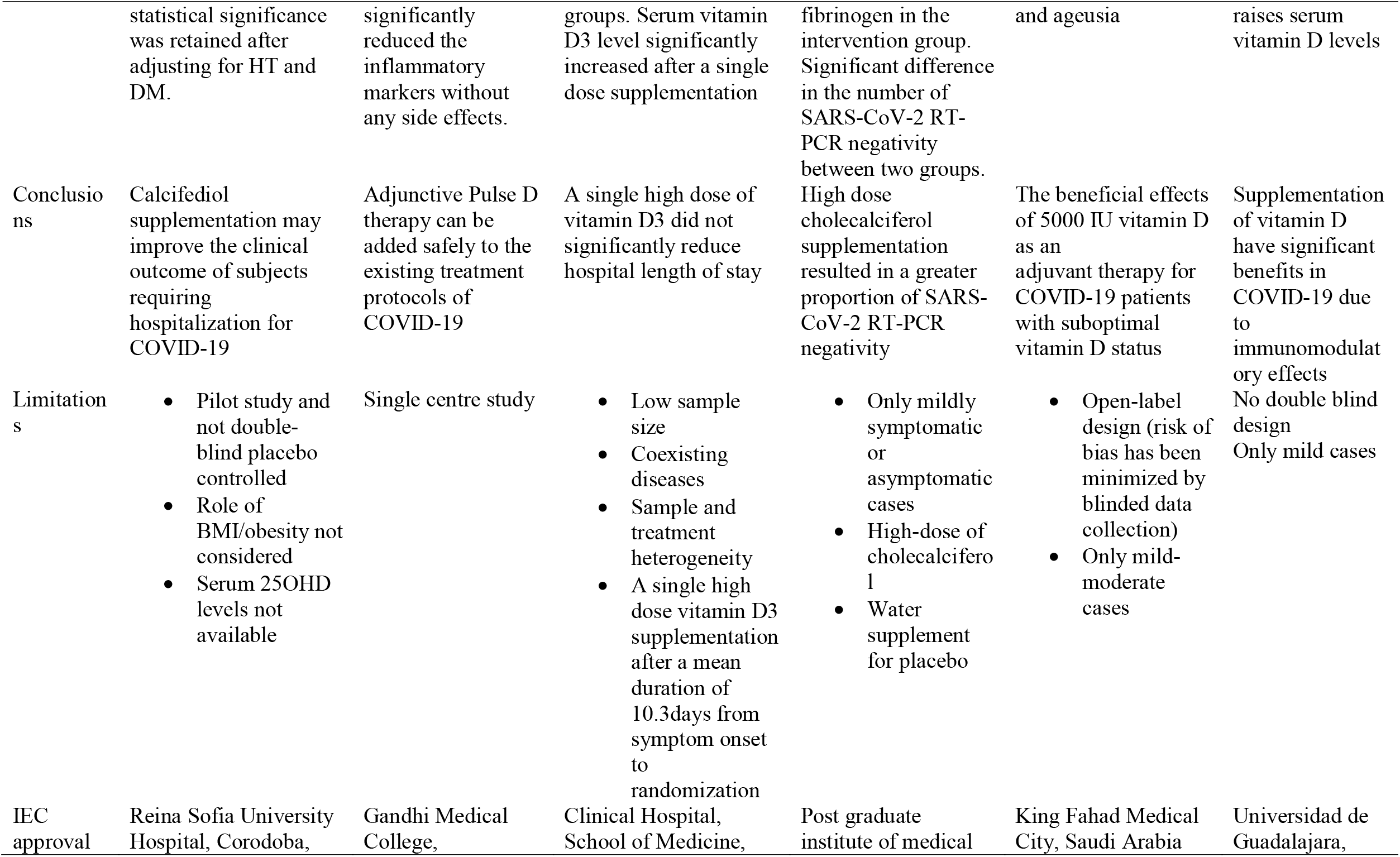
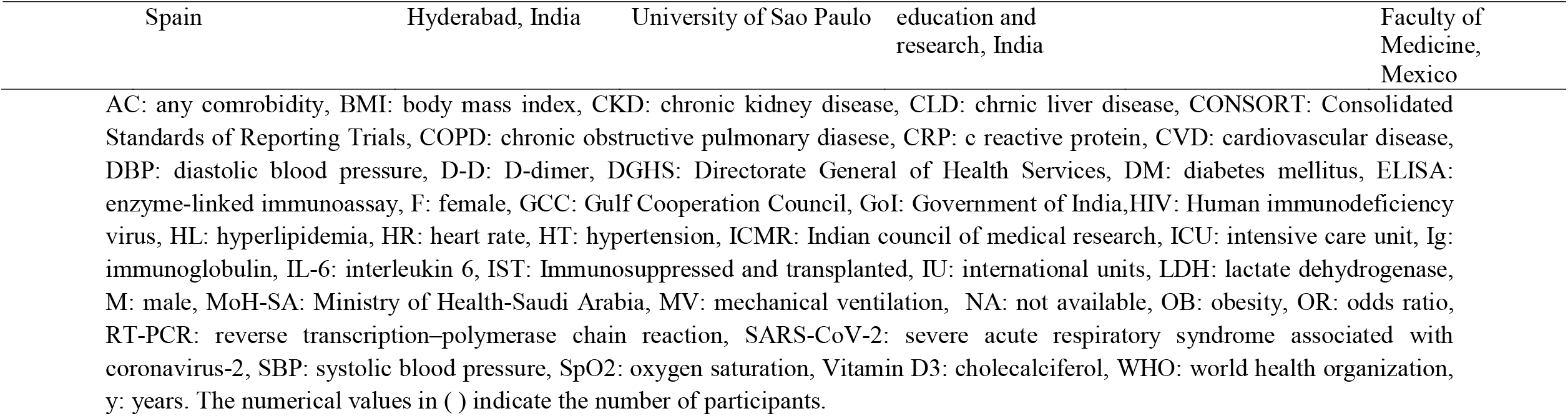
The study characteristics of all RCTs included in the meta-analysis

There were two multi-center [16,18] and four single center RCTs [14,15,17,19], one double-blinded [16] and four registered clinical trials [14–16,18]. Vitamin D treatment was compared to placebo in two studies [16,17], non-vitamin control in three studies [14,15,19], and standard treatment comparator group in one study [18]. While Castillo et al. [14] used calcifediol with an allocation ratio of 2:1; all other studies used cholecalciferol with an allocation ratio of 1:1. The baseline vitamin D statuses in three studies [15,17,18] were reported to be sub-optimal and one study reported a separate outcome analysis in vitamin D deficient participants [16]. The vitamin D sufficiency status, treatment doses, follow-up durations, adverse events and study limitations are detailed in Table 1. The risk of bias assessment based on five domains and the overall bias of included RCTs is presented the supplementary appendix.

The collective evidence in Fig.2 shows that vitamin D treatment was significantly associated with reduced risk of COVID-19 severity when six observations on the number events for symptom severity, ICU care and mechanical ventilation were pooled (RR = 0.46, 95% CI 0.23 to 0.93, Z=2.16, p=0.03, I^2^ = 52%). But the pooled estimate from four studies showed that the use of vitamin D was not significantly associated with ICU outcome alone (RR = 0.11, 95% CI 0.15 to 1.30, Z=1.48, p=0.14, I^2^ = 66%).

**Fig.2.**
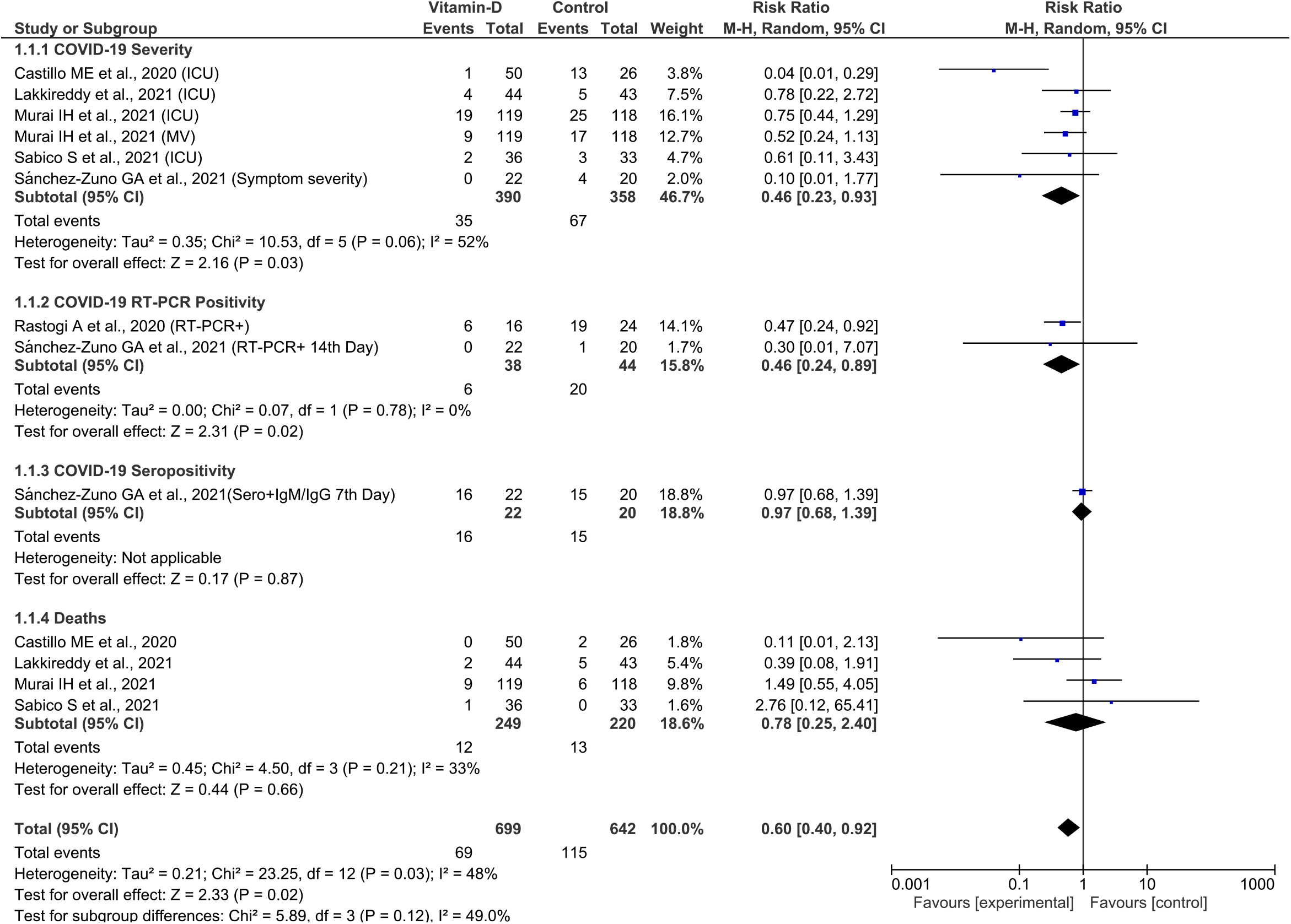
The Forest plot for association of vitamin D intervention in COVID-19

The pooled estimate from two studies showed a statistically significant RR for COVID-19 RT-PCR positivity (RR = 0.46, 95% CI 0.24 to 0.89, Z=2.31, p=0.02, I^2^ = 0%). Whereas the pooled evidence from four studies showed that the association of vitamin D with mortality outcome was not statistically significant (RR = 0.78, 95% CI 0.25 to 2.40, Z=0.66, p=0.02, I^2^ = 33%). However, when all the observations on all reported outcomes were pooled, there was statistically significant evidence on the use of vitamin D treatment in reducing overall COVID-19 related outcomes (RR = 0.60, 95% CI 0.40 to 0.92, Z=2.33, p=0.02, I^2^ = 48%). The test for subgroup differences was not statistically significant (I^2^ = 49%, p = 0.12).

The results of sub-group analysis were presented in Table 2. None of the outcomes in different categories of subgroups showed statistically significant RR values. No statistically significant difference was observed for the pooled estimate of outcomes from studies with vitamin D suboptimal status. The sensitivity analysis performed leaving-out any one of the included trials at a time and repeating the analysis showed statistically non-significant RR values for individual outcomes. Whereas, for all studied outcomes together, the pooled RR remained statistically significant after leaving our any particular study/observation. The I^2^ value significantly changed from 48% to 5% after leaving-out a study by Castillo et al. (ICU and mortality observations) and repeating the analysis suggestive of major source of heterogeneity. The funnel plot analysis (Fig.3) with Begg’s (p = 0.17) and Egger tests (p = 0.14) on all the outcomes across all the RCTs indicated no significant publication bias.

**Table 2.**
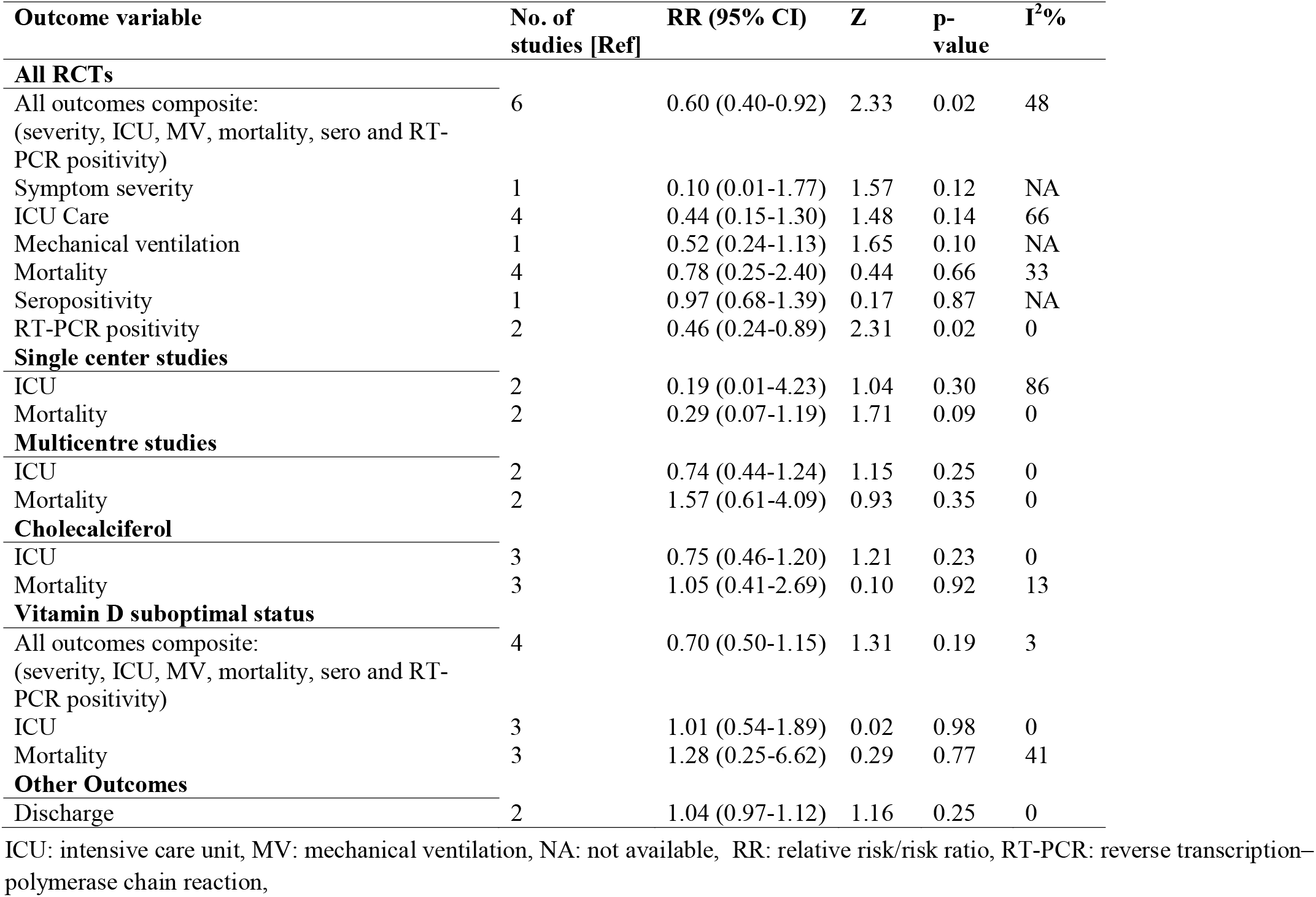
The results of subgroup analysis

| **Outcome variable** | **No. of studies [Ref]** | **RR (95% CI)** | **Z** | **p-value** | **I<sup>2</sup>%** |
| --- | --- | --- | --- | --- | --- |
| **All RCTs** |  |  |  |  |  |
| All outcomes composite:<br>(severity, ICU, MV, mortality, sero and RT-PCR positivity) | 6 | 0.60 (0.40-0.92) | 2.33 | 0.02 | 48 |
| Symptom severity | 1 | 0.10 (0.01-1.77) | 1.57 | 0.12 | NA |
| ICU Care | 4 | 0.44 (0.15-1.30) | 1.48 | 0.14 | 66 |
| Mechanical ventilation | 1 | 0.52 (0.24-1.13) | 1.65 | 0.10 | NA |
| Mortality | 4 | 0.78 (0.25-2.40) | 0.44 | 0.66 | 33 |
| Seropositivity | 1 | 0.97 (0.68-1.39) | 0.17 | 0.87 | NA |
| RT-PCR positivity | 2 | 0.46 (0.24-0.89) | 2.31 | 0.02 | 0 |
| **Single center studies** |  |  |  |  |  |
| ICU | 2 | 0.19 (0.01-4.23) | 1.04 | 0.30 | 86 |
| Mortality | 2 | 0.29 (0.07-1.19) | 1.71 | 0.09 | 0 |
| **Multicentre studies** |  |  |  |  |  |
| ICU | 2 | 0.74 (0.44-1.24) | 1.15 | 0.25 | 0 |
| Mortality | 2 | 1.57 (0.61-4.09) | 0.93 | 0.35 | 0 |
| **Cholecalciferol** |  |  |  |  |  |
| ICU | 3 | 0.75 (0.46-1.20) | 1.21 | 0.23 | 0 |
| Mortality | 3 | 1.05 (0.41-2.69) | 0.10 | 0.92 | 13 |
| **Vitamin D suboptimal status** |  |  |  |  |  |
| All outcomes composite:<br>(severity, ICU, MV, mortality, sero and RT-PCR positivity) | 4 | 0.70 (0.50-1.15) | 1.31 | 0.19 | 3 |
| ICU | 3 | 1.01 (0.54-1.89) | 0.02 | 0.98 | 0 |
| Mortality | 3 | 1.28 (0.25-6.62) | 0.29 | 0.77 | 41 |
| **Other Outcomes** |  |  |  |  |  |
| Discharge | 2 | 1.04 (0.97-1.12) | 1.16 | 0.25 | 0 |

ICU: intensive care unit, MV: mechanical ventilation, NA: not available, RR: relative risk/risk ratio, RT-PCR: reverse transcription–polymerase chain reaction,

**Fig.3.**
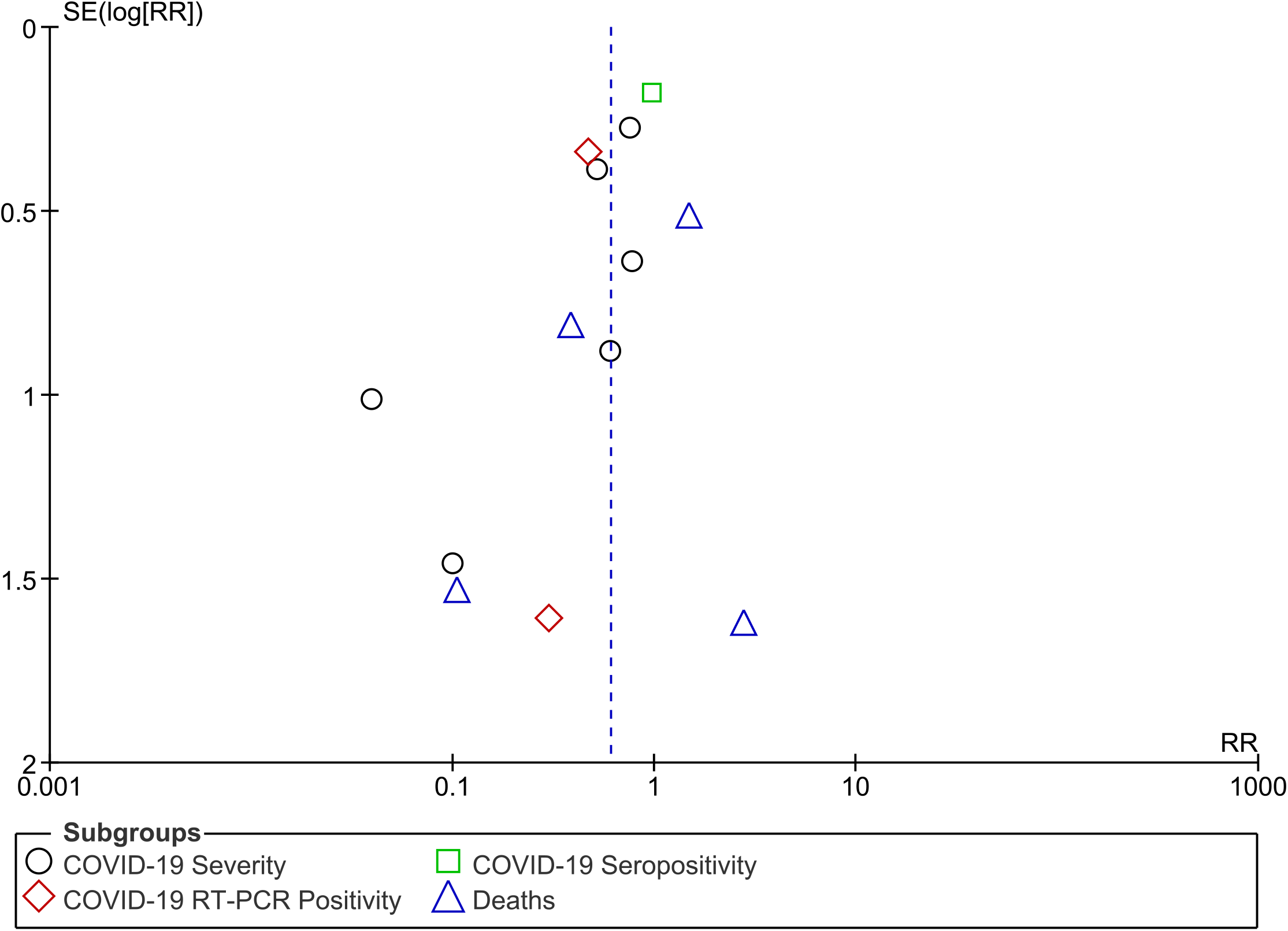
The Funnel plot for publication bias

## Discussion

This meta-analysis of RCTs showed that COVID-19 patients supplemented with vitamin D had reduced overall risk for all outcomes. The collective overall evidence on severity, ICU care, mortality, sero and RT-PCR positivity events reported in all trials indicated that COVID-19 patients treated with vitamin D showed lower rates of these outcomes relative to patients receiving no-vitamin D/standard/placebo. Though there were no statistically significant differences in the individual outcomes of ICU admission and mortality, the respective RRs indicated a decrease in the rates of these outcomes in vitamin D treated groups. However, there was a statistically significant decrease in the rates of RT-PCR positivity in COVID-19 patients supplemented with vitamin D.

The first multicenter double-blind RCT study by Murai et al. [16] enrolled 237 moderate-severe COVID-19 patients. It had 119 patients in the experimental group treated with a single high dose of vitamin D_3_ (200000 IU orally) and 118 patients in the placebo group receiving peanut oil. The results to do not support the use of a high dose of vitamin D as it did not significantly reduce the length of hospital stay, hospital discharge, ICU admission and rates of mechanical ventilation and mortality. Similar findings were reported in subgroups of patients (57 in intervention and 58 in placebo arms) with vitamin D deficiency at baseline (<20 ng/mL), despite of achieving sufficient status (≥30 ng/mL) in 86.7% of the vitamin D3 group post intervention. This study reports more mortality events in the intervention arm (9/119) than the placebo (6/118) group.

In another multicenter RCT [18] randomizing 73 mild-moderate COVID-19 patients with suboptimal vitamin D status into experimental (n=36) and standard-comparator (n=33) groups receiving 5000 IU and 1000 IU of oral cholecalciferol daily for two weeks. This study though reports a significantly shorter recovery time to symptoms (even after adjusting for age, sex, BMI and D-dimer) in the intervention arm, no significant differences in ICU, mortality events and days to discharge were reported between groups. This study differs from that of Murai et al. [16] as it excludes severe COVID-19 cases, vitamin D dosage and duration, using standard comparator group in place of placebo, and in defining the suboptimal vitamin D status (<50 nmol/L). Further, this study also differs from all other trials as 47% of randomized participants had also received vitamin C supplements. The significant increase in vitamin D levels reported in treatment arm (5000 IU) post intervention along with other study findings are to be interpreted with caution to the baseline vitamin D levels in the comparator arm. The post-treatment vitamin D levels (62.5 nmol/L) of the intervention arm are similar to that of the pre-treatment levels (63 nmol/L) in comparator arm (p=0.67).

In an RCT by Sánchez-Zuno et al. [19] 42 mild COVID-19 patients were randomized to intervention arm (22 cases receiving 10000 IU of vitamin D_3_ orally for 14 days) and comparator arm that receives no vitamin D3 (n=20). A stratified analysis based on the sufficient (≥30 ng/mL) and insufficient (<30 ng/mL) baseline vitamin status indicated a significantly increased number of COVID-19 symptoms in the later group (p=0.03). It was found that the intervention arm had significantly increased vitamin D levels post-treatment and presented fewer symptom severities on the seventh and fourteenth day of follow-up. The intervention arm also had lesser rates of seropositivity and RT-PCR positivity on the seventh and fourteenth day, respectively. In a study by Rastogi et al. [17] a similar observation was reported with a significant decrease in the proportion of SARS-CoV-2 RNA negativity in the intervention arm before day-21 (p<0.01). This study randomized 40 mildly symptomatic or symptomatic COVID-19 patients into intervention and placebo arms of 20 cases each. The intervention arm received 60000 IU of cholecalciferol daily for 7 days and continued for another 7 days in six cases (who did not achieve a therapeutic target of >50 ng/mL on day 7) and distilled water was supplied to placebo group.

There were two open label RCTs [14,15]. Lakkireddy et al. [15] randomized 130 mild-moderate COVID-19 cases, of which 87 cases who completed the study were analyzed in the intervention (n=44) and comparator (n=43) groups. The intervention arm received 60000 IU of oral vitamin D3 daily for 8-10 days and the outcomes were recorded till 21 days. Supplementation resulted in a significant increase in vitamin D levels with a lower rate of ICU and mortalities in the intervention arm as compared to the comparator group. In the only trial using calcifediol, Castillo et al. [14] randomized 76 patients into intervention (n=50) and comparator (n=26) groups depending on whether or not supplemented with calcifediol. The oral calcifediol supplemented varied at admission (0.532 mg), on days 3 and 7 (0.266 mg) and weekly until ICU/discharge (0.266 mg). This study concludes that vitamin D treatment resulted in significantly less probability of ICU admission and the statistical significance retained even after adjusting for comorbidities like diabetes and hypertension. However, there is no information available on the baseline and post-treatment vitamin D levels.

In general, it has been demonstrated that vitamin D induce antimicrobial peptides and mediates antiviral, apoptotic and autophagic activities [12,23]. The protective immuno-modulatory effects of this fat-soluble steroid vitamin have been reported in respiratory diseases [24,25]. Studies have proposed vitamin D deficiency as leading candidate in association with COVID-19 susceptibility, severity and progression [26,27]. However, there is no strong evidence through RCTs on the therapeutic benefits of vitamin D supplementation in COVID-19 outcomes. Our study results based on the available RCTs are suggestive of the overall beneficial effect of vitamin D treatment when all the observations across all RCTs were pooled as an overall effect size. Though no statistically significant differences were observed for ICU care and mortality outcomes individually, the observed RR values are suggestive of decrease in the rate of these outcomes in vitamin D treated COVID-19 patients. This meta-analysis based on RCTs is first of its kind on the subject and the results are supportive of vitamin D use in COVID-19. Further, as there is compatible evidence in the form of a meta-analysis of observational studies on the use of vitamin D in COVID-19 [13], the results of this study strongly suggests the need for future/ongoing RCTs to consider better designs, large sample sizes adequate enough to assess the effect of vitamin D supplementation on the individual COVID-19 related outcomes.

However, this study has some limitations. First, the heterogeneity observed in the meta-analysis could be due to methodological, participant and treatment variations of the included trials. While the single center RCTs have mainly contributed to the heterogeneity, leaving-out a study by Castillo et al. [14] decreased the I^2^ values from 48 to 5%, 66 to 0%, and 33 to 13% for the overall outcome pooling the results of all RCTs, ICU and mortality outcomes, respectively. This open label trial differs from all other RCTs as it uses calcifediol in varied concentrations at different time periods of the study. Second, there are only two placebo-controlled trials, one double-blinded study that uses a single high dose of vitamin D. Third, although no significant loss to the follow-up were reported in the RCTs, the proportion of participants and the criteria for sufficient and deficient vitamin D status varied across the trials. Fourth, the variations in the COVID-19 severity, comorbidities proportions and standard care treatment strategies could have influenced the heterogeneity and the overall result. Finally, the difference in the study settings, timings, randomization, blinding, and data collection strategies could have influenced the outcomes. None of the trials reported any adverse events due to vitamin supplementation. As there are only two and three trials respectively in the years 2020 [14,17] and 2021 [15,16,18,19] including small sample sizes, this meta-analysis strongly recommends for more RCTs for better evaluating the role of vitamin D in COVID-19 patients. Therefore, the evidence obtained upon completion of several ongoing trials [28] (CORONAVIT, COVITD-19, COVIDIOL, VIVID and COVIT-TRIAL) will be crucial in better determination on vitamin D in association with COVID-19.

In conclusion, vitamin D use was associated with significant decrease in rates of COVID-19 related events when all the outcomes were pooled across all RCTs. However, there was no significant difference observed for the relative risk for ICU admission and mortality outcomes upon vitamin D supplementation. The overall pooled results in addition to a significant decrease in the rates of RT-PCR positivity observed in this study are suggestive of the possible beneficial effects of vitamin D. These results would indicate the need for more RCTs in supportive of ongoing trials evaluating the effect of vitamin D in COVID-19.

## Supporting information

Supplementary Appendix: Risk of bias assessment

## Data Availability

All the data relevant to this article is available with the corresponding author and can be shared upon receipt of a request on Email.

## Author Statement

S.R.V: Conceptualization, literature search, data extraction, methodology, analysis, interpretation, writing, supervising, reviewing and editing. B.T: literature search, assistance in data extraction, analysis, review and writing. H.R: conduction and verification of literature search, data extraction results and analysis.

## Acknowledgements

Dr. S.R. Varikasuvu specially acknowledge Bhairavi Sisters (Sahasra and Aagneya) for the time I could not give them during this work, as it was performed before 8 am and after 6 pm from its initiation to completion.

## Figure legends

**Supplementary Appendix**: Risk of bias assessment

