## Supplementary Appendix: Risk of bias assessment for "COVID-19 and Vitamin D (Co-VIVID Study): a systematic review and meta-analysis of randomized controlled trials"

Risk of bias assessment was done using the Cochrane risk of bias tool.

**Ref:**

RoB 2: a revised tool for assessing risk of bias in randomised trials. DOI: [10.1136/bmj.l4898](https://doi.org/10.1136/bmj.l4898)

<https://pubmed.ncbi.nlm.nih.gov/31462531/>

The risk of bias was assessed at five domains (D1-D5 as stated in the below figure) for all the included RCTs.

Our assessment indicated some concerns of bias in all the studies. Although we could not judge any bias/concerns in a study by Murai et al. This interpretation should be cautioned to the use of single high dose of vitamin D3 supplementation in the concerned study.

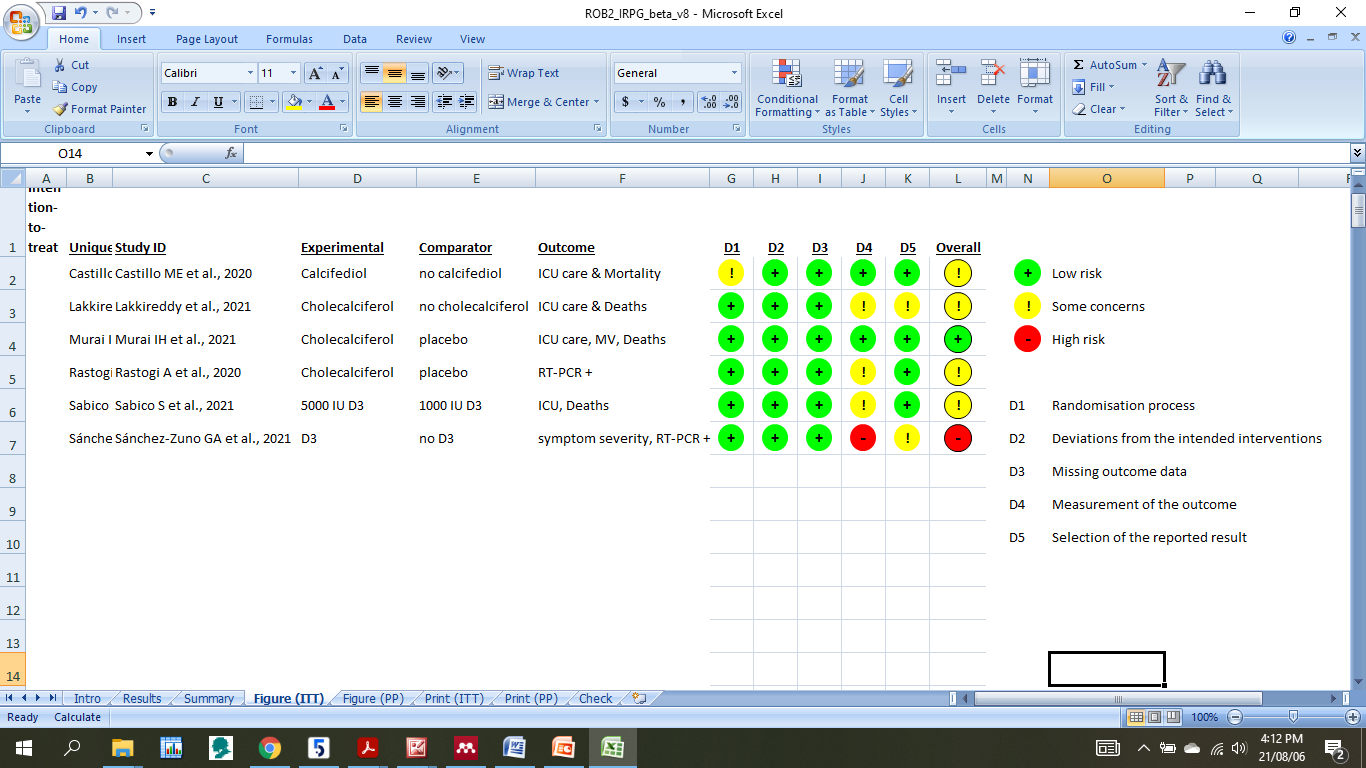

The below figure indicates domain level bias for all the included studies along with the overall bias. As seen in the figure, the overall bias of the included studies could be judged as having “some concerns” related to randomization, blinding, outcome analysis and reporting of results.

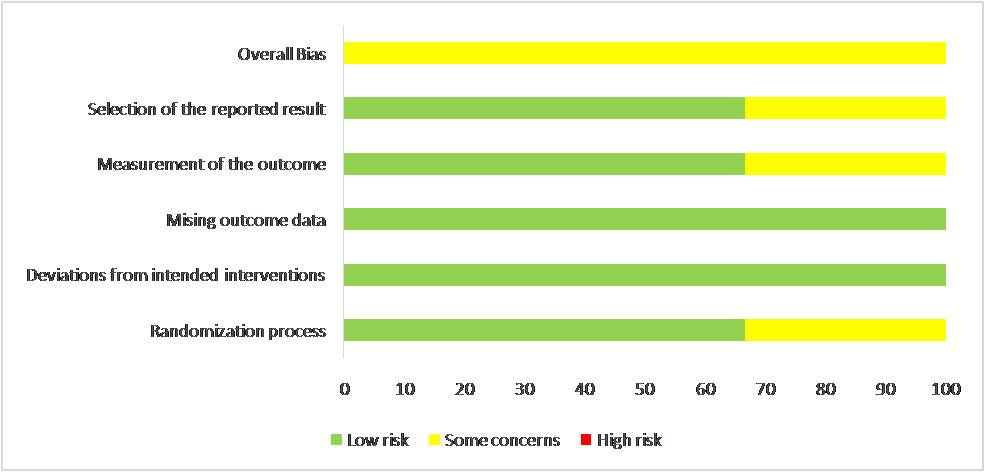
